# eHEALS.com.br: Automating Digital Health Literacy Assessment and Personalized Feedback in Brazil

**DOI:** 10.64898/2026.01.22.26344614

**Authors:** Thalles Cotta Fontainha, Vera Maria Benjamim Werneck, Claudia Cappelli

## Abstract

This paper describes eHEALS.com.br, a web-based platform that automates the administration of the Brazilian eHealth Literacy Scale (eHEALS-Br). The system collects responses online, scores users in real time, and provides personalized feedback based on five levels of digital health literacy. A systematic literature review was conducted to map existing instruments and identify gaps related to automation, temporal control, and inclusion. The platform architecture combines a React and TypeScript frontend with a Node.js and MongoDB backend, deployed on a secure virtual private server. In a pilot study with 12 participants, the automated eHEALS-Br scale demonstrated excellent internal consistency (Cronbach’s alpha = 0.929), and additional questionnaire dimensions also showed good reliability. The results demonstrate the feasibility of automating eHEALS-Br, supporting both population studies and individualized clinical use, and reinforcing the potential of digital health tools for monitoring and improving health literacy in Brazil.

## I. Introduction

The term “e-Saúde” in Portuguese or “ehealth” (Electronic Health), referred to health practices that were facilitated or improved by the use of digital technologies [1], [2]. In 2006, Cameron D. Norman and Harvey A. Skinner introduced eheals, an instrument designed to measure “ehealth literacy” manually. This instrument aims to evaluate users’ ability to understand and apply health information obtained through digital resources [3]. eHeals has gained recognition worldwide for its effectiveness and versatility over the years, becoming a standard in research related to digital health literacy.

Some other tools were globally recognized, applied, and validated. Prominent examples included HLS-14 (Health Literacy Scale) in Japan, which had 14 questions dealt with on the form *“Q14”* of its acronym. These tools were applied to understand how individuals accessed, understood, and utilized health information in clinical and public health contexts, demonstrating proper reliability and validity [4]. Another important example is the European HLS-Q47, a 47-item instrument used to assess comprehensive health literacy in various aspects, such as healthcare, disease prevention, and health promotion [5]. Although reduced versions such as HLS-EU-Q16 and HLS-EU-Q6 had been validated in many countries, including Italy and Sweden [6], [7], such instruments were inadequate for an automated system in the Brazilian context due to factors such as: first, they had been granted to manual application, limiting their application, Integration in digital platforms; Second, they needed the modular structure necessary to provide real-time feedback; Third, they did not incorporate the specificities of the health system or the sociocultural characteristics of a population as large and diverse as the Brazilian.

The platform eheals.com.br represents an innovation that not only automates eHEALS-Br but also integrates features combining rankings between surveyed groups, custom feed-back, and post-end analyses, thereby expanding its practical usefulness. This scalability enables the tool to be applied to various scenarios, ranging from population research to individualized clinical care, thereby meeting the needs of both public and private health sectors. In addition, the structured data generated by the platform has great potential to feed scientific production, offering valuable insights for the development of new strategies and interventions in the field of health literacy. The creation of the platform also addresses the need to make the evaluation of health literacy (LS in Portuguese) more inclusive and accessible. The implementation of eHEALS-Br in an automated format not only preserves the analytical essence of the original instrument but also significantly expands its applicability. This innovation simplifies user interaction with the tool, providing feedback and scores attributed to each user in the research through an automated evaluation algorithm, and optimizes large-scale data collection and analysis.

The relevance of this project is further reinforced by its alignment with the United Nations Sustainable Development Goals (SDGs) [8], which are illustrated in the slogan in Figure 1. The platform directly contributes to ODS 3 (*Health and Well-being*) by facilitating access to evaluation instruments that can enhance quality of life and promote preventive health. It relates to ODS 4 (*Quality Education*) by democratizing access to educational health tools and promoting digital health literacy. Finally, it meets ODS 9 (*Industry, Innovation and Infrastructure*) when developing technological solutions for complex public health challenges. This convergence with ODS reinforces the platform’s transformative potential, positioning it as a strategic tool for sustainable development in the health sector.

**Fig. 1.**
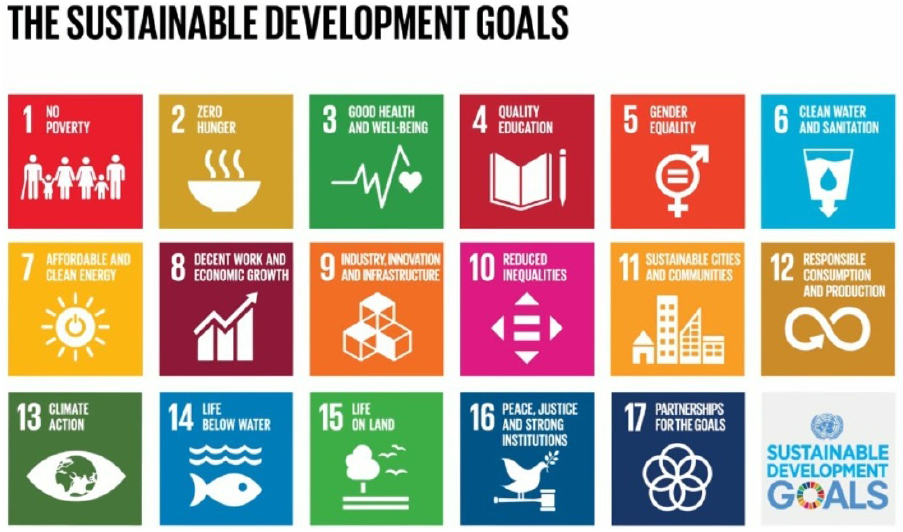
The 17 Sustainable Development Goals (SDGs) established by the United Nations [8].

### A. Conceptual Framework

This section aims to present conceptual and methodological definitions and justifications that expand the understanding of the proposal of this work. The main concepts related to digital health, platform structure eheals.com.br, and methodological decisions adopted throughout the study are discussed.

The platform eheals.com.br Structure your analysis in four conceptual pillars: “Evaluation” (interface usability perceptions) [9], “Agreement” (adhesion to functional benefits) [10] “Frequency” (Informational Consumer Patterns) [11], and “Capacity” (domain of digital health skills) [12]. These dimensions translate the concept of digital literacy, as measured by eHEALS-Br [13], into practice by integrating individual evaluations with health information systems [14].

## II. Related Work

### A. Literature Review

The initial search strategy retrieved records from four databases using the selected English terms. Table I presents the raw results, specifying the number of records identified for each search term in SciELO, PubMed, IEEE, and ACM, respectively.

The prism diagram in Fig. 2 shows the filtering and selection process for systematic mapping. This process begins with identification and concludes with the inclusion of relevant studies [15]. Records were gathered from four databases, totaling 1,490. Eight duplicate records were removed before screening. Automation tools discarded 1,361 ineligible records, and an additional 8 were removed for non-health-related reasons.

**TABLE I.** Results of the research of the terms in English.

| Flag | Research term | SciELO | PubMed | IEEE | ACM | Total |
| --- | --- | --- | --- | --- | --- | --- |
| A1 | eHEALS | 11 | 220 | 1 | 12 | 244 |
| A2 | Health Literacy Scale | 14 | 401 | 0 | 5 | 420 |
| A3 | Literacy Measurement | 1 | 90 | 4 | 6 | 101 |
| A4 | eHealth Literacy | 20 | 669 | 17 | 19 | 725 |
| **Total** | **A1 + A2 + A3 + A4** | **46** | **1380** | **22** | **42** | **1490** |

Note: Research data after PRISMA.

**Fig. 2.**
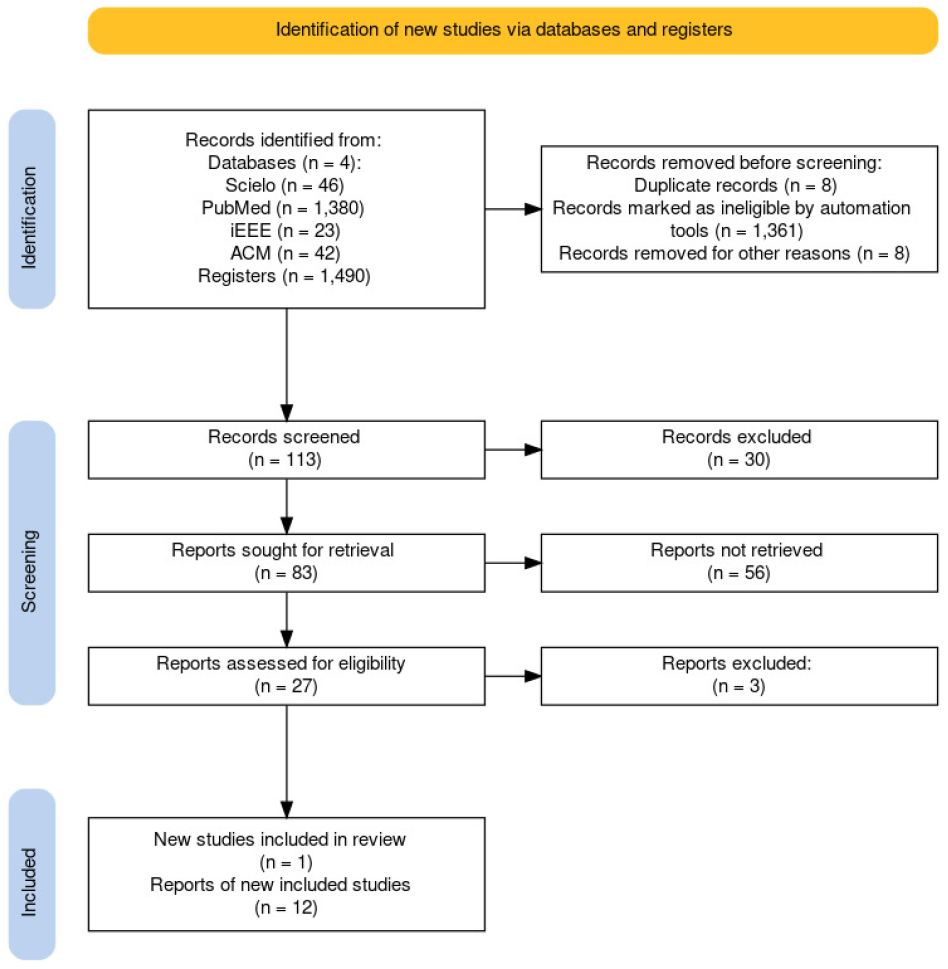
PRISMA Flow Diagram [15]^1^.

Table II details the progression of articles through each filtering phase, displaying how many papers were retained at each stage from all four data sources.

**TABLE II.**
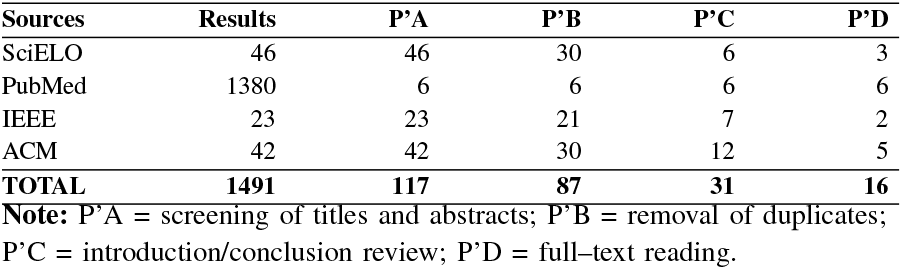
Selection of Articles.

| Sources | Results | P'A | P'B | P'C | P'D |
| --- | --- | --- | --- | --- | --- |
| SciELO | 46 | 46 | 30 | 6 | 3 |
| PubMed | 1380 | 6 | 6 | 6 | 6 |
| IEEE | 23 | 23 | 21 | 7 | 2 |
| ACM | 42 | 42 | 30 | 12 | 5 |
| **TOTAL** | **1491** | **117** | **87** | **31** | **16** |

Note: P'A = screening of titles and abstracts; P'B = removal of duplicates; P'C = introduction/conclusion review; P'D = full-text reading.

### B. Key Findings from the Literature

This result is based on the article by Norman and Skinner, authors of the foundational eHEALS tool. While their article is not available in the consulted databases, it is widely recognized and frequently cited in digital health literacy literature. Thus, it is appropriate to acknowledge Norman and Skinner’s pioneering contributions as seen in their work [3], which highlights the importance and application of eHEALS in digital health literacy assessments. eHEALS, created and validated to assess digital health literacy among young people in the United States and Canada, underwent sequential data collection and statistical analysis. Results indicate eHEALS is a reliable, unidimensional tool, making it effective for measuring digital health literacy.

The standardization of the eHEALS scale scores was analyzed by Mialhe et al. (2023) [17] in a cross-sectional study of 502 Brazilian adults. These participants completed the eHEALS instrument and a sociodemographic questionnaire. The analysis used decision trees and discriminant analysis. The study found that education has a significant impact on the scale’s results, suggesting a need to control digital health literacy classifications by participants’ educational level. Based on this validated Brazilian translation, with formal authorization from both the national team and the original authors, the following study was conducted.

Figures 3 and 4 summarize the findings from the systematic review. Figure 3 presents the worldwide distribution of the included studies. There is a concentration of publications in North American and European countries. In addition, there is a category classified as “International,” which represents multicenter investigations. Figure 4, in turn, displays the correlation matrix between the coded variables of the analyzed studies. For example, there is a negative correlation between *year* and *country encoded*, suggesting that countries with a greater presence of studies tended to present older publications. There is also a negative association between *population encoded* and *instrument encoded*, indicating differences in the choice of instruments according to the evaluated population profile.

**Fig. 3.**
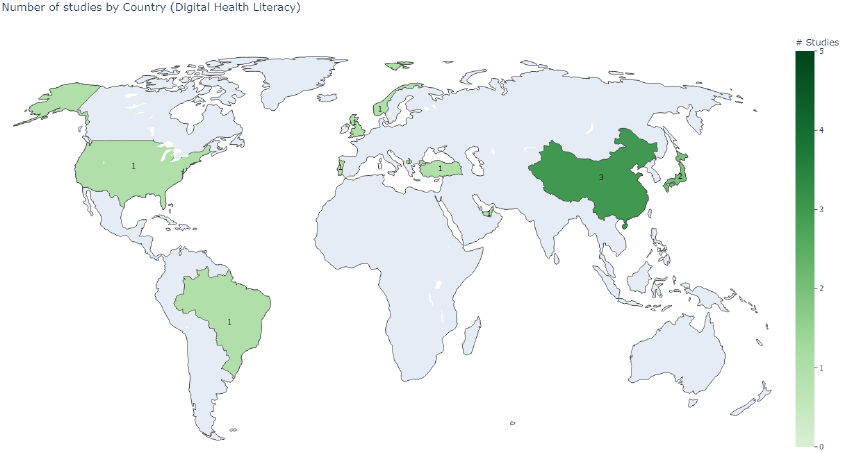
Global distribution of included studies on digital health literacy. The map displays the number of studies by country, with a green scale indicating increasing intensity based on the number of publications. The uniform light green color represents the scope of studies classified as “International.”

**Fig. 4.**
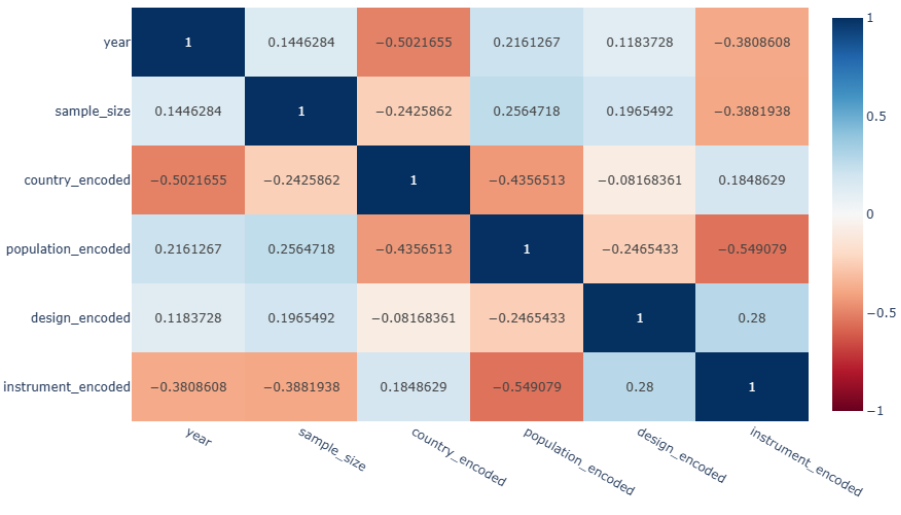
Correlation matrix between coded variables of the studies on digital health literacy. Color intensity indicates the strength and direction of the Pearson correlation (**red = negative, blue = positive**). A negative correlation is observed between *year* and *country encoded*, suggesting that countries with more studies tended to present older publications, and between *population encoded* and *instrument encoded*.

### C. The eHEALS-Br Automated Platform

The systematic literature review, as shown by [18] and [2] in Table II, reveals three main gaps in applying eHEALS. First, the lack of automated platforms with temporal control of an Information System (all analyzed studies use traditional methods of application); second, the absence of mechanisms for comparison among participants (no study proposes systems for comparative ranking); and third, the need for adaptations to promote digital inclusion (as highlighted by [19] and [20]). The platform eheals.com.br addresses these challenges through: (i) automation of the eHEALS-Br data collection process with controlled collection periods; (ii) implementation of an accessible design with simplified language; and (iii) generation of comparative rankings at the end of each cycle. As a result, participants gain clear insight into both their own scores and how they compare to others, which can provide motivation, encourage engagement, and promote learning. The solution follows the Brazilian Ministry of Health’s guidelines [14]. It balances methodological rigor by keeping validated psychometric properties and increases digital accessibility by overcoming barriers found by [19]. This supports the growing computerization of Brazil’s health system.

The validated Brazilian version of eHEALS, as described by [17], preserves the original psychometric structure [3] while incorporating specific linguistic adaptations, such as the revision of the term *“health resources”* to better fit the Brazilian context. This robust methodological basis, although limited to traditional applications, supports the innovation introduced by the automated platform.

## III. Methods

After the literature review, the *snowball sampling* method was adopted, as described in prior studies [21], [22]. This non-probabilistic technique is particularly suitable for studies with hard-to-reach populations. The process began with 12 participants in the municipality of Rio de Janeiro. The strategy enabled organic expansion to other municipalities and states outside Rio de Janeiro through chain referrals - a fundamental characteristic of the method. As predicted, the approach combined initial direct dissemination with snowball expansion [22]. This combination proved efficient for ensuring a consistent sample base, reducing operational costs, and capturing unplanned diversity. The results confirm the technique’s adequacy for studies in digital health literacy.

Data collection took place between March 17 and 25, 2025, via the eheals.com.br platform and Google Forms. These tools enabled the standardization of sociodemographic data collection and the multidimensional assessment of digital literacy. The process included mandatory response validation features and Likert scale options. This approach optimized quantitative analysis. The final sample was diverse in terms of professional profiles and digital literacy levels. This diversity demonstrated the method’s effectiveness in digital contexts, where connections between participants transcend geographical boundaries. The division of the study questions into two specific categories, *Evaluation* and *Agreement*, follows methodological principles established in the specialized literature. Questions classified as *Evaluation* address participants’ qualitative perceptions regarding aspects such as clarity, ease, and security, whereas items grouped under *Agreement* involve the explicit acceptance of statements or perceptions of the platform’s usefulness. This conceptual separation is consistent with the recommendations of [23] and the psychometric criteria described in [24], which emphasize the importance of grouping conceptually similar items to enhance the interpretative coherence and analytical consistency of the results.

### A. System Overview (eHEALS-Br Platform)

The manual eHEALS-Br instrument was adapted into an interactive digital interface that uses clear textual language and multimedia content to improve the user experience when answering the eight eHEALS-Br questions. The platform maintains the eight validated questions from Brazil and is accessible on multiple devices. Visual guides and GIF instructions were added to help users. The next subsections describe the platform’s main modules: automated evaluation, data security, and end-of-survey feedback.

#### 1) Automated Evaluation Algorithm

Algorithms were implemented to process the responses of eHEALS-Br in real time, generating individual analyses on the users’ LS with a database structured on a web platform. The algorithms provided a qualitative interpretation of the results, explaining what a specific score meant and which areas needed improvement. Note: Comparative analyses and group creation were performed by the researcher in version 1.1.3 of the platform^2^. The system has versioning for future updates, allowing managers to register surveys and other functions.

#### 2) Data Security

The security of user data was guaranteed as a priority, with the implementation of robust security measures, such as SSL certificates and secure hosting, such as VPS. The platform adhered to the highest data security standards, ensuring the protection and privacy of user information.

#### 3) Feedback at the end of a Survey

The platform provided clear feedback to the user about what their scores meant and how they could improve, using simple and accessible language to ensure understanding by people with different educational levels.

#### 4) Accessible Design and Communication Integration

In the textual dimension, the principles of Plain Language were applied in accordance with international guidelines [25], with the adaptation of the original paper layout to a dynamic web interface that incorporates animated visual elements on the platform. This combined approach, visual and textual, aimed to preserve the technical accuracy of the original instrument and its translation, while enhancing the accessibility and autonomy of users on the platform. The results of this accessibility verification are illustrated in Figure 5, which presents a practical analysis of the interface’s compliance with key accessibility requirements.

**Fig. 5.**
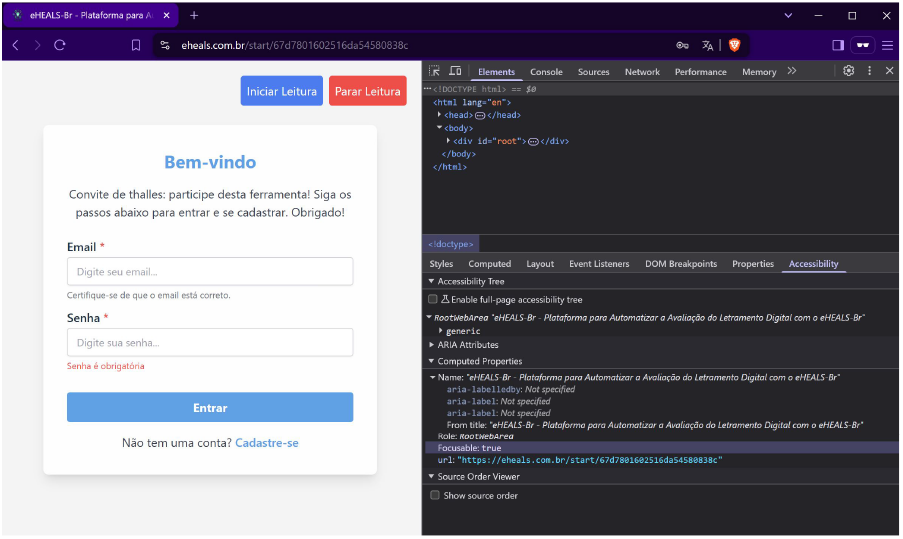
Accessibility analysis of the eHEALS-Br platform interface showing compliance with WCAG 2.1 guidelines.

The digital platform of eHEALS-Br was developed with a modern architecture, utilizing React and TypeScript for the frontend to provide a responsive and accessible interface, and Node.js with MongoDB for the backend to efficiently manage questionnaire data. The infrastructure, hosted on a secure VPS (virtual private server), was subjected to rigorous technical validation tests, including load and compatibility, ensuring robustness. The process included the formal validation of the original instrument’s holders, ensuring the integral preservation of its eight items, the Likert scale, and its psychometric properties in the Brazilian version.

The automation transformed the static questionnaire into an interactive quiz, with a participation flow that goes from access through a unique link to the final personalized feedback. The platform incorporates accessibility guidelines (WCAG 2.1) and features such as continuous saving, vocalization of questions, and visualization adapted for mobile devices, thereby maintaining the scientific validity of the original instrument while leveraging the advantages of digitalization, including secure storage and automated data analysis.

### B. Scoring and Reliability

The rating system adopted on the eheals.com.br platform was based on cumulative scoring, as per the eHEALS-Br instrument, where each response has a specific weight not disclosed to participants. This preventive strategy sought to avoid social desirability bias - the tendency to select responses perceived as more favorable. The scoring scale ranged from 8 points (all responses scored 1) to 40 points (all responses scored 5).

The Likert scale, used exclusively in the original instrument, was a five-point scale used for each item, ranging from (1) Strongly disagree, (2) Somewhat disagree, (3) Unsure, (4) Somewhat agree, and (5) Strongly agree. The rating algorithm was programmed to resolve potential ties by following a hierarchy: primary criterion of total score, secondary criterion of participant age (with older respondents prioritized), and, as a final criterion, the date of response submission (with older respondents prioritized).

### C. Study Design & Sampling

Data collection employed the *snowball sampling* method, initiated with direct dissemination via WhatsApp to three health professionals (initial controls) who forwarded the questionnaire to their networks, resulting in 12 complete respondents eligible for statistical analysis in the present work. This approach allowed: (i) organic reach within the municipality of Rio de Janeiro; (ii) reduction of homophily bias by the controlled profile of the initial participants [21]; and (iii) operational efficiency, obtaining qualified responses with minimal investment in dissemination [22].

Data collection took place between March 17 and 25, 2025, during which specific strategies for quality control were implemented. The platform incorporated validation algorithms that identified and filtered incomplete or duplicate responses through checking IP addresses and cookies. All procedures are strictly adhered to in accordance with ethical guidelines, with a digital consent form integrated into the registration process (CAAE: 81347724.2.0000.5282).

### D. Study Period & Participant Classification

At the end of the established period (March 17-25, 2025), the 12 eligible participants completed the evaluation on the automated platform. The results were classified into five categories (see Table III) based on the total eHEALS-Br score: 1st group (36 to 40 points), 2nd group (30 to 35 points), 3rd group (24 to 29 points), 4th group (16 to 23 points), and 5th group (8 to 15 points). This classification into five groups was created in the present work.

**TABLE III.**
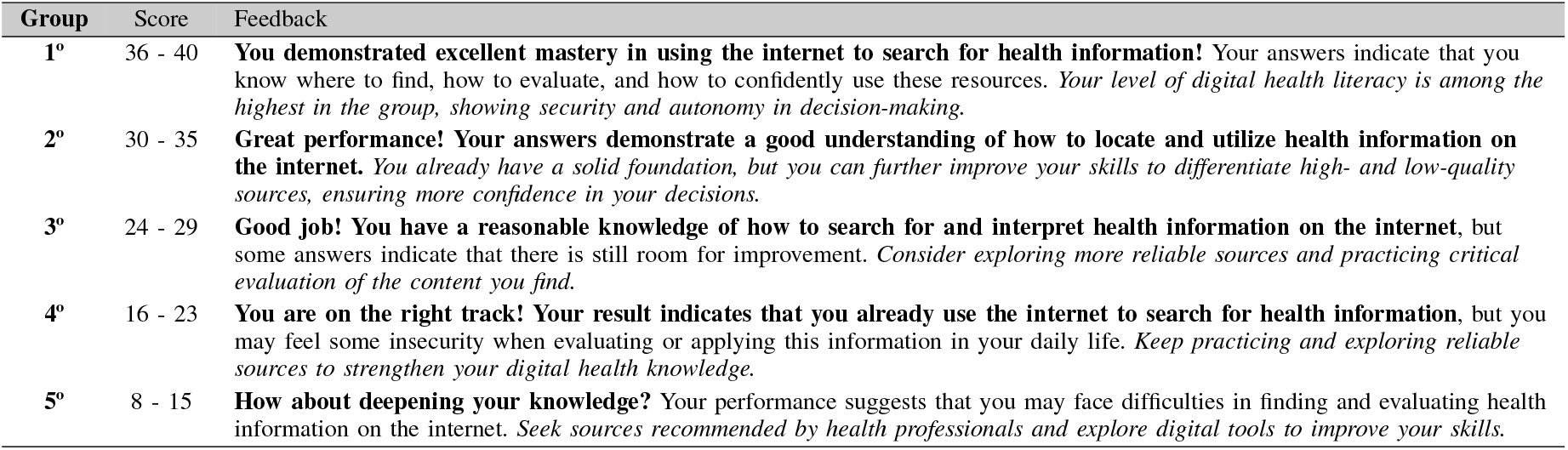
Platform classification and feedback.

| Group | Score | Feedback |
| --- | --- | --- |
| 1° | 36 - 40 | **You demonstrated excellent mastery in using the internet to search for health information!** Your answers indicate that you know where to find, how to evaluate, and how to confidently use these resources. *Your level of digital health literacy is among the highest in the group, showing security and autonomy in decision-making.* |
| 2° | 30 - 35 | **Great performance! Your answers demonstrate a good understanding of how to locate and utilize health information on the internet.** *You already have a solid foundation, but you can further improve your skills to differentiate high- and low-quality sources, ensuring more confidence in your decisions.* |
| 3° | 24 - 29 | **Good job! You have a reasonable knowledge of how to search for and interpret health information on the internet,** but some answers indicate that there is still room for improvement. *Consider exploring more reliable sources and practicing critical evaluation of the content you find.* |
| 4° | 16 - 23 | **You are on the right track! Your result indicates that you already use the internet to search for health information,** but you may feel some insecurity when evaluating or applying this information in your daily life. *Keep practicing and exploring reliable sources to strengthen your digital health knowledge.* |
| 5° | 8 - 15 | **How about deepening your knowledge?** Your performance suggests that you may face difficulties in finding and evaluating health information on the internet. *Seek sources recommended by health professionals and explore digital tools to improve your skills.* |

This stratification allowed a direct comparative analysis of the participants’ performance in relation to digital health literacy. For example, if the first place in a group scored 15 points, although leading that group, they would receive feedback for group 5, which suggests improvements to the user.

The classification criterion was based on the total score obtained by summing the weights of each question. Each question ranged from 1 to 5 points, with a minimum score of 8 and a maximum of 40. In the event of a tie, which did not occur in this study, tie-breaking criteria would be applied: total score, respondent age, and, if necessary, the most recent response date. The platform’s approach, eheals.com.br, aimed to provide feedback and comparative metrics within the group, utilizing the platform’s resources.

The feedback presented in Table III was developed in the present work. It was based on the following digital health literacy profiles for the platform eHEALS.com.br^3^. For the groups with higher scores, the feedback was inspired by [20], highlighting confidence and critical thinking. In the intermediate groups, common difficulties identified by [19] were incorporated, such as evaluating the credibility of sources. The text for the group with the lowest performance was based on [18] and [26], adopting a motivational tone with practical guidelines. All feedback maintains recognition of effort, clarity in observed skills, and accessible recommendations, using simple language.

The five groups in Table III(8-15, 16-23, 24-29, 30-35, 36-40) organize the evolution of competencies in digital health literacy. They represent stages from initial search to critical analysis. The groupings are based on theoretical references to offer guidance adapted to each stage of development.

### E. Post-Cycle Feedback Mechanism

The automated feedback system aimed to be effective by providing participants with a clear assessment of their performance, combining their scores with a personalized analysis based on the classification in Table III. This approach allowed participants to understand their ranking in the group and identify both their competencies and areas for development, even without knowing the actual sample size. The detailed responses automatically generated by the platform eheals.com.br ultimately provided specific and accessible guidance for each proficiency level, as demonstrated above.

## IV. Results

The initial questions (Q01-Q12) from the Google Forms are used solely for registration and sociodemographic identification of research participants. These are excluded from the qualitative analysis of user experience for the eheals.com.br platform. Instead, the analysis specifically targets responses from Q13 onward, which examine core dimensions of user experience. The following table IV presents results for key aspects: Ease of Access (Q13: Accessibility and Audio Controls), Clarity of Questions (Q15: Inclusion and Clarity), Platform Usefulness (Q16: Website Usefulness), Difficulties of Understanding (Q14: Difficulties of Understanding), Perception of Safety (Q21: Safety), and Confidence in Literacy (Q28: Health Literacy). This approach provides insight into both platform usability and the overall user experience, as well as the effectiveness of the tool’s communication.

**TABLE IV.** Methodological categories assigned to questions Q13 - Q29.

| Question | Question Summary | Category |
| --- | --- | --- |
| Q13 | Accessibility and Audio Controls | Evaluation |
| Q14 | Difficulty of Understanding and Language | Evaluation |
| Q15 | Inclusion and Clarity of Questions | Evaluation |
| Q16 | Did you find the site useful? | Agreement |
| Q17 | Ease of Access | Evaluation |
| Q18 | Clarity of Questions | Evaluation |
| Q19 | Inclusion (all ages) | Agreement |
| Q20 | Knowledge Improvement | Agreement |
| Q21 | Security | Evaluation |
| Q22 | Usefulness of Feedback | Agreement |
| Q23 | Traditional Literacy (internet use freq.) | Frequency |
| Q24 | Scientific Literacy | Capacity |
| Q25 | Information Literacy (perceived quality) | Evaluation |
| Q26 | Media Literacy | Frequency |
| Q27 | Computational Literacy (tool familiarity) | Evaluation |
| Q28 | Health Literacy (in online decisions) | Capacity |

Note: Prepared by the author (2025), based on Google Forms data.

### A. Instruments & Conceptual Categories

The study was designed based on the methodological principles consolidated by [23] and [24], with the aim of structuring the qualitative responses obtained in the study into two distinct conceptual categories: Evaluation and Agreement. This division reflects essential aspects of the collected perceptions. The “Evaluation” category captures participants’ qualitative judgments about the platform’s ease of use, clarity, and security, while “Agreement” addresses the degree of users’ explicit agreement regarding the usefulness and acceptance of certain features or functionalities of the tool. This qualitative-quantitative mapping facilitates statistical analysis, enabling a coherent interpretation of the data and enhancing subsequent reliability, as presented in the next section.

To better adapt to the study, two new categories were added to the Likert scale: “Frequency”, which measures how regularly participants access digital health resources, and “Capacity”, which assesses their self-perception of technical skills. These additions preserve the essential properties of the original Likert scale while expanding its applicability to the digital health context, following approaches validated in previous work [27].

Table IV defines categories covering responses to questions Q13-Q29. The “Evaluation” category includes users’ perceptions of ease, clarity, security, and familiarity with the platform (Q13, Q15, Q17, Q18, Q21, Q25, Q27). The “Agreement” category covers respondents’ explicit agreement on perceived usefulness, knowledge improvement, inclusion for different age groups, confidence, and capacity in scientific and health literacy (Q16, Q19, Q20, Q22, Q24, Q28). Other questions measure frequency (Q23, Q26), difficulty (Q14), and general comments (Q29), which are outside these categories.

The 15 questions analyzed in the questionnaire (Q13-Q28) were categorized into four distinct conceptual categories: “Evaluation”, “Agreement”, “Frequency” and “Capacity”. This organization (Table V) is based on the eHEALS-Br structure adapted from [18]. This classification enabled the analysis of different aspects of users’ perceptions - from judgments about the platform’s usability to their digital interaction patterns, degree of agreement with its perceived usefulness, and digital health competencies.

**TABLE V.**
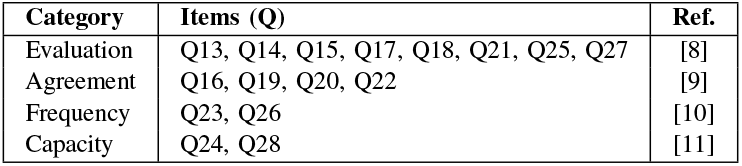
Distribution of questions by conceptual category and theoretical basis.

| Category | Items (Q) | Ref. |
| --- | --- | --- |
| Evaluation | Q13, Q14, Q15, Q17, Q18, Q21, Q25, Q27 | [8] |
| Agreement | Q16, Q19, Q20, Q22 | [9] |
| Frequency | Q23, Q26 | [10] |
| Capacity | Q24, Q28 | [11] |

The categorization adopted was inspired by the theoretical contributions of the analyzed articles. “Evaluation” encompassed judgments about the functional and experiential aspects of the platform, including accessibility, linguistic clarity, and perceived quality [28]. The “Agreement’ analyzed adherence to propositions about instrumental value, ranging from immediate utility to impacts on learning [29]. “Frequency” describes patterns of interaction with digital health ecosystems [30], including the intensity of use and the critical processing of information. “Capacity” diagnosed cognitive and technical competencies for effective engagement with digital health literacy resources [12].

Table VI summarizes the content of each quantitative item. It specifies the assessed dimension and provides the complete wording of each question. This structured description clarifies how variables were operationalized for the analyses. It ensures transparency in measuring usability, comprehension, and digital health literacy across the questionnaire’s 15 items.

**TABLE VI.** Detailed description of study variables.

| Question | Dimension | Full Description |
| --- | --- | --- |
| Q13 | Usability | Did you find it easy to access health information on the platform? |
| Q14 | Language | Did you have any difficulty understanding the questions, and do you think simpler language would help? |
| Q15 | Clarity | Did the eHEALS-Br questions seem clear to you? |
| Q16 | Usefulness | Did you find the eheals.com.br website useful? |
| Q17 | Navigation | Subjective assessment of ease of navigation on the platform |
| Q18 | Design | Perception of visual clarity and graphic organization of the content |
| Q19 | Inclusion | Do you think people of all ages can easily use the platform? |
| Q20 | Learning | Did the information provided help improve your knowledge about digital health? |
| Q21 | Safety | Did you feel safe using the eheals.com.br platform? |
| Q22 | Feedback | How useful was the feedback requested after interacting with the website? |
| Q23 | Frequency | How often do you use the internet to search for health information? |
| Q24 | Scientific Literacy | Do you feel able to understand and apply the scientific information you find online? |
| Q25 | Quality | How do you evaluate the quality of the health information you find online? |
| Q26 | Criticism | Can you interpret and critique health-related media content? |
| Q27 | Digital Skills | How familiar are you with using digital tools to access health information? |
| Q28 | Autonomy | How confident are you in making health decisions based on information found online? |

Note: Adapted from Google Forms (2025) from the eheals.com.br platform.

### B. Quantitative Results

Table VII presents the consolidated results of the 8 questions applied on the platform, with the 12 respondents distributed according to their perceptions (N = 12). Additionally, Table VIII details the responses to the 16 Google Forms questions.

**TABLE VII.** Platform answers from e heals.com.br for the 8 e heals-br questions (V1 - V8), N = 12.

| Platform Answers × Questions |  |  |  |  |  |  |  |
| --- | --- | --- | --- | --- | --- | --- | --- |
| V1 | V2 | V3 | V4 | V5 | V6 | V7 | V8 |
| 5 | 5 | 4 | 5 | 5 | 5 | 5 | 5 |
| 4 | 4 | 4 | 4 | 4 | 3 | 3 | 3 |
| 4 | 4 | 4 | 4 | 4 | 4 | 4 | 4 |
| 4 | 4 | 4 | 4 | 4 | 4 | 4 | 4 |
| 3 | 4 | 4 | 3 | 5 | 2 | 5 | 2 |
| 4 | 4 | 4 | 4 | 3 | 4 | 3 | 4 |
| 2 | 3 | 4 | 1 | 1 | 1 | 4 | 4 |
| 5 | 4 | 4 | 5 | 4 | 5 | 4 | 4 |
| 5 | 5 | 5 | 5 | 5 | 4 | 5 | 4 |
| 5 | 4 | 4 | 4 | 3 | 3 | 4 | 5 |
| 1 | 1 | 1 | 3 | 2 | 1 | 2 | 1 |
| 1 | 2 | 3 | 4 | 3 | 2 | 1 | 3 |

Notes:
- V1: I know what health resources are available on the internet. - V2: I know where to find useful health resources on the internet. - V3: I know how to find useful health resources on the internet. - V4: I know how to use the internet to answer my health questions. - V5: I know how to use the health information I find on the internet to help me. - V6: I have the skills to evaluate the health resources I find on the internet. - V7: I can differentiate between high-quality and low-quality health resources. - V8: I feel confident using internet information to make health decisions. - Scale: 1) Strongly disagree 2) Somewhat disagree 3) Unsure 4) Somewhat agree 5) Strongly agree.

**TABLE VIII.**
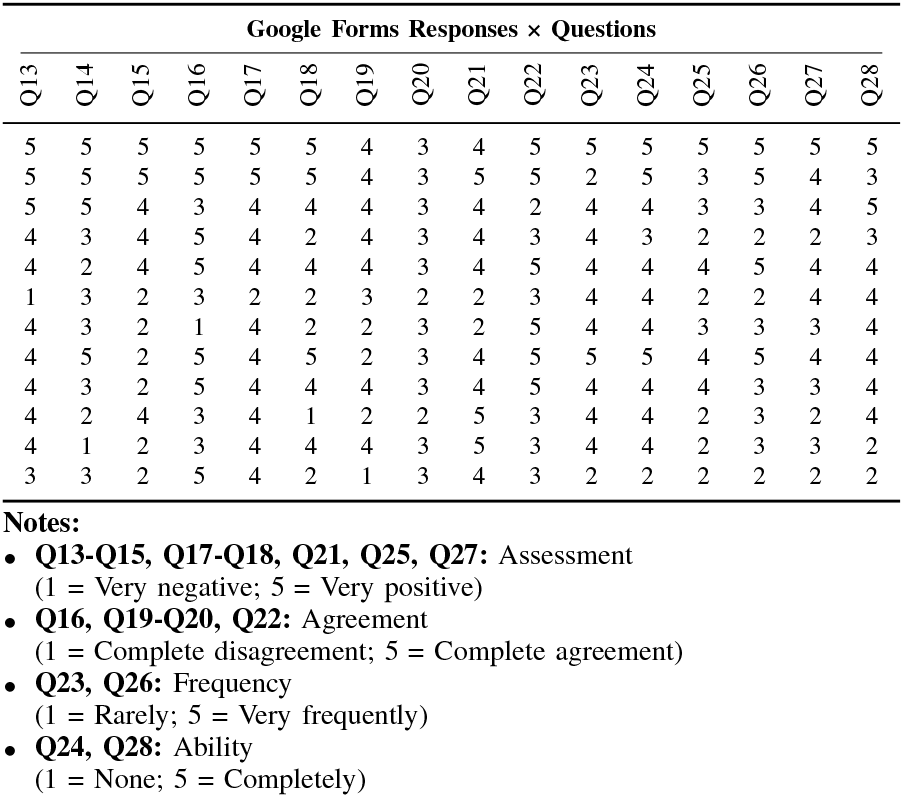
Google Forms responses - 16 questions (Q13-Q28), ***N* = 12**.

| Google Forms Responses × Questions |  |  |  |  |  |  |  |  |  |  |  |  |  |  |  |
| --- | --- | --- | --- | --- | --- | --- | --- | --- | --- | --- | --- | --- | --- | --- | --- |
| Q13 | Q14 | Q15 | Q16 | Q17 | Q18 | Q19 | Q20 | Q21 | Q22 | Q23 | Q24 | Q25 | Q26 | Q27 | Q28 |
| 5 | 5 | 5 | 5 | 5 | 5 | 4 | 3 | 4 | 5 | 5 | 5 | 5 | 5 | 5 | 5 |
| 5 | 5 | 5 | 5 | 5 | 5 | 4 | 3 | 5 | 5 | 2 | 5 | 3 | 5 | 4 | 3 |
| 5 | 5 | 4 | 3 | 4 | 4 | 4 | 3 | 4 | 2 | 4 | 4 | 3 | 3 | 4 | 5 |
| 4 | 3 | 4 | 5 | 4 | 2 | 4 | 3 | 4 | 3 | 4 | 3 | 2 | 2 | 2 | 3 |
| 4 | 2 | 4 | 5 | 4 | 4 | 4 | 3 | 4 | 5 | 4 | 4 | 4 | 5 | 4 | 4 |
| 1 | 3 | 2 | 3 | 2 | 2 | 3 | 2 | 2 | 3 | 4 | 4 | 2 | 2 | 4 | 4 |
| 4 | 3 | 2 | 1 | 4 | 2 | 2 | 3 | 2 | 5 | 4 | 4 | 3 | 3 | 3 | 4 |
| 4 | 5 | 2 | 5 | 4 | 5 | 2 | 3 | 4 | 5 | 5 | 5 | 4 | 5 | 4 | 4 |
| 4 | 3 | 2 | 5 | 4 | 4 | 4 | 3 | 4 | 5 | 4 | 4 | 4 | 3 | 3 | 4 |
| 4 | 2 | 4 | 3 | 4 | 1 | 2 | 2 | 5 | 3 | 4 | 4 | 2 | 3 | 2 | 4 |
| 4 | 1 | 2 | 3 | 4 | 4 | 4 | 3 | 5 | 3 | 4 | 4 | 2 | 3 | 3 | 2 |
| 3 | 3 | 2 | 5 | 4 | 2 | 1 | 3 | 4 | 3 | 2 | 2 | 2 | 2 | 2 | 2 |

Notes:
- Q13-Q15, Q17-Q18, Q21, Q25, Q27: Assessment (1 = Very negative; 5 = Very positive) - Q16, Q19-Q20, Q22: Agreement (1 = Complete disagreement; 5 = Complete agreement) - Q23, Q26: Frequency (1 = Rarely; 5 = Very frequently) - Q24, Q28: Ability (1 = None; 5 = Completely)

### C. Reliability Analysis

The internal consistency of the eheals.com.br platform scale (items V1 to V8 of Table VII) was assessed using Cronbach’s

Alpha coefficient and a Confidence Interval (CI) of *α* = 0.929; 95% CI [0.847 - 0.976], showing excellent reliability according to the criteria established by [31]. At the same time, the scale applied via Google Forms (items Q13 to Q28) showed *α* = 0.891 (95% CI [0.815 - 0.976]), indicating good internal consistency, as shown in Table IX.

**TABLE IX.** Interpretation of reliability coefficients.

| Alpha ($\alpha$) | Interpretation |
| --- | --- |
| $\alpha > 0.90$ | Excellent reliability (observed in response eheals.com.br) |
| $0.80 \leq \alpha \leq 0.90$ | Good reliability (Google Forms case) |
| $0.70 \leq \alpha < 0.80$ | Acceptable |
| $\alpha < 0.70$ | Problematic |

Note: Interpretation of reliability coefficients according to Nunnally [31].

**TABLE X.** Reliability Coefficients by Category.

| Category (Items) | Alpha ($\alpha$) |
| --- | --- |
| Full Item Set (Q13-Q28) | 0.891 |
| Assessment (Q13,Q14,Q15,Q17,Q18,Q21,Q25) | 0.836 |
| Agreement (Q16,Q19,Q20,Q22) | 0.505 |
| Frequency (Q23,Q26) | 0.351 |
| Ability (Q24,Q27,Q28) | 0.833 |

The results suggest that both tools demonstrate adequate internal consistency for assessing digital health literacy. However, when directly comparing the two, the eheals.com.br platform achieved excellence, surpassing the other tool. The analysis was performed using the pingouin package in Python 3.11, following robust statistical protocols for estimating confidence intervals.

## V. Discussion

### A. Relationship Between Response Categories and Assessed Dimensions

The questionnaire responses were converted into quantitative variables using four categories: Evaluation, Agreement, Frequency, and Capability. However, the statistical reliability analysis focused on three main psychometric dimensions: Evaluation, Usefulness, and Capability. The distinction exists because the initial categories are based on language type in the questions, such as whether they are affirmative or agreement-based. In contrast, the dimensions reflect what is being measured, like perceived usability, practical application (usefulness), or digital self-efficacy (capability).

Therefore, questions in the “Agreement” category might be part of either the “Evaluation” or “Usefulness” dimension, depending on their purpose. Separating these helps ensure the analysis aligns with eHEALS-Br definitions and remains consistent with earlier studies.

### B. Comparative Reliability Analysis of Instruments

The eight questions administered on the eheals.com.br platform were analyzed as a single scale, respecting the unidimensional model of the internationally validated eHEALS-Br instrument. This decision is justified both by the original structure of the scale and by the excellent internal consistency observed (*α* = 0,929), which indicates high reliability for measuring digital health literacy. The questionnaire administered via Google Forms (Q13-Q28) was analyzed multidimensionally, with the items grouped into four categories. The complete set showed good internal consistency (*α* = 0,891), and the categories Assessment (*α* = 0,836) and Capacity (*α* = 0,833) demonstrated satisfactory indices.

This weakness suggests that, although users recognize the practical purposes of the platform, there may be inconsistencies in how they interpret its usefulness and frequency of use, possibly influenced by different user profiles or varying levels of familiarity with the topics covered [18], [19], [26].

### C. Study Limitations

The study has two main limitations: (1) the transformation of qualitative responses into a Likert scale, although methodologically valid for parametric analyses, implies the simplification of the nuances of individual perceptions, assuming equidistant intervals between response categories [27]; (2) the multidimensional analysis of the questionnaire administered via Google Forms revealed considerable variations in the reliability coefficients between categories-ranging from *α* = 0.351 (Frequency) to *α* = 0.836 (Evaluation), which indicates inconsistencies in the internal coherence of some dimensions. These results suggest that certain categories, such as Agreement and Frequency, may require reformulation of the items to achieve greater psychometric robustness in future studies.

### D. Practical Impact and Academic Contribution

The source code and related materials are available at the author’s GitHub repository.^4^. This work highlights the importance of developing master’s research that bridges theory and practice through automation and real-world implementation. The project exemplifies how academic investigations can generate tangible tools that optimize evaluation processes and promote innovation in digital health literacy assessment in Brazil.

## VI. Conclusion

The findings indicate that the eHEALS-Br instrument demonstrated strong methodological consistency, particularly within the “*Assessment*” (*α* = 0.836) and “*Capacity*” (*α* = 0.833) dimensions. These results suggest that participants exhibited a robust understanding of both the platform’s usability and their capacity to utilize online health information. The five-level scoring system (8 - 40 points) effectively distinguished users according to their digital health literacy, with intermediate scores occurring most frequently.

In contrast, the “*Agreement*” (*α* = 0.505) and “*Frequency*” (*α* = 0.351) dimensions exhibited lower reliability, which may be attributable to ambiguous wording or inconsistencies in item interpretation. Future revisions should prioritize rephrasing or restructuring these items to improve psychometric robustness.

Conducted as part of a Master’s dissertation in the Professional Graduate Program in Telemedicine and Digital Health at the State University of Rio de Janeiro (UERJ), this study demonstrates the feasibility of automating the eHEALS-Br instrument and highlights the significance of applied research within Brazil’s digital health literacy landscape. Broader, multi-regional validations are recommended to ensure that *eheals*.*com*.*br* remains a reliable and equitable tool across Brazil’s diverse social and cultural contexts.

## Data Availability

All data produced in the present study are available upon reasonable request to the authors.

## Acknowledgment

The author extends special thanks to his advisors, Prof. Vera Maria Benjamim Werneck and Prof. Claudia Cappelli, for their guidance and support, which was provided with patience. Their essential assistance began during the master’s program and continues to this day. The author also acknowledges the faculty and colleagues of the Professional Master’s Program in Telehealth and Digital Health (UERJ) – CNPq Research Group and his current Ph.D. studies in the Graduate Program in Instrumentation and Applied Optics (PPGIO/CEFET-RJ).^5^. Their contributions have greatly enriched his interdisciplinary training in health and technology. Finally, he expresses heartfelt appreciation to his wife, Ana Cláudia Rodrigues Fontainha.

This study was financed in part by the Coordenação de Aperfeiçoamento de Pessoal de Nível Superior - Brasil (CAPES) - Finance Code 001.

## Thalles Cotta Fontainha

earned a B.Sc. in Computer Science from the State University of Rio de Janeiro (UERJ) in 2022 and an M.Sc. in Telemedicine and Telehealth from UERJ in 2025. He is currently pursuing a Ph.D. in Instrumentation and Applied Optics at the Federal Center for Technological Education of Rio de Janeiro (CEFET/RJ). His research focuses on machine learning applications in digital health, with emphasis on 3D convolutional neural networks for knee MRI analysis and the automation of the eHEALS-Br platform. He also has experience in the development and integration of health information systems. He is a member of the Brazilian Health Literacy Network (REBRALS) and the Brazilian Computer Society, and received the Best Short Paper Award in 2025.

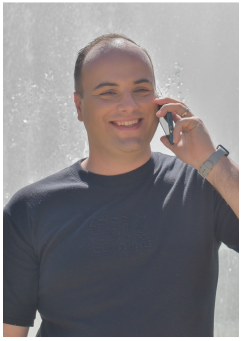

## Vera Maria Benjamim Werneck

is a Full Professor in the Department of Informatics and Computer Science at the State University of Rio de Janeiro (UERJ), where she coordinates the Professional Master’s Program in Telemedicine and Telehealth. She holds a Doctorate and Master’s degree in Systems and Computer Engineering, as well as a Bachelor’s degree in Mathematics with a specialization in Informatics, all from the Federal University of Rio de Janeiro (UFRJ). Her areas of expertise include software engineering, requirements engineering, medical systems, and digital health. Her current research focuses on telemedicine, plain language, and health informatics applications in public health. She is a member of the Brazilian Computer Society (SBC) and the Brazilian Telemedicine and Telehealth Association (ABTms).

## Claudia Cappelli

is a Professor of Computer Science at the State University of Rio de Janeiro (UERJ), where she teaches in the Professional Master’s Program in Telemedicine and Telehealth and collaborates with the Graduate Program in Information Systems at the Federal University of Rio de Janeiro (UFRJ). She earned a Ph.D. in Informatics from PUC-Rio (2009), an M.Sc. in Information Systems from UFRJ (2000), and a B.Sc. in Informatics from UERJ (1985), and completed postdoctoral research at UNIRIO (2010) and UFRJ (2020). Her professional experience includes process management roles at Citibank and Telemar. Her research interests encompass business process management, enterprise architecture, digital governance, plain language.

---

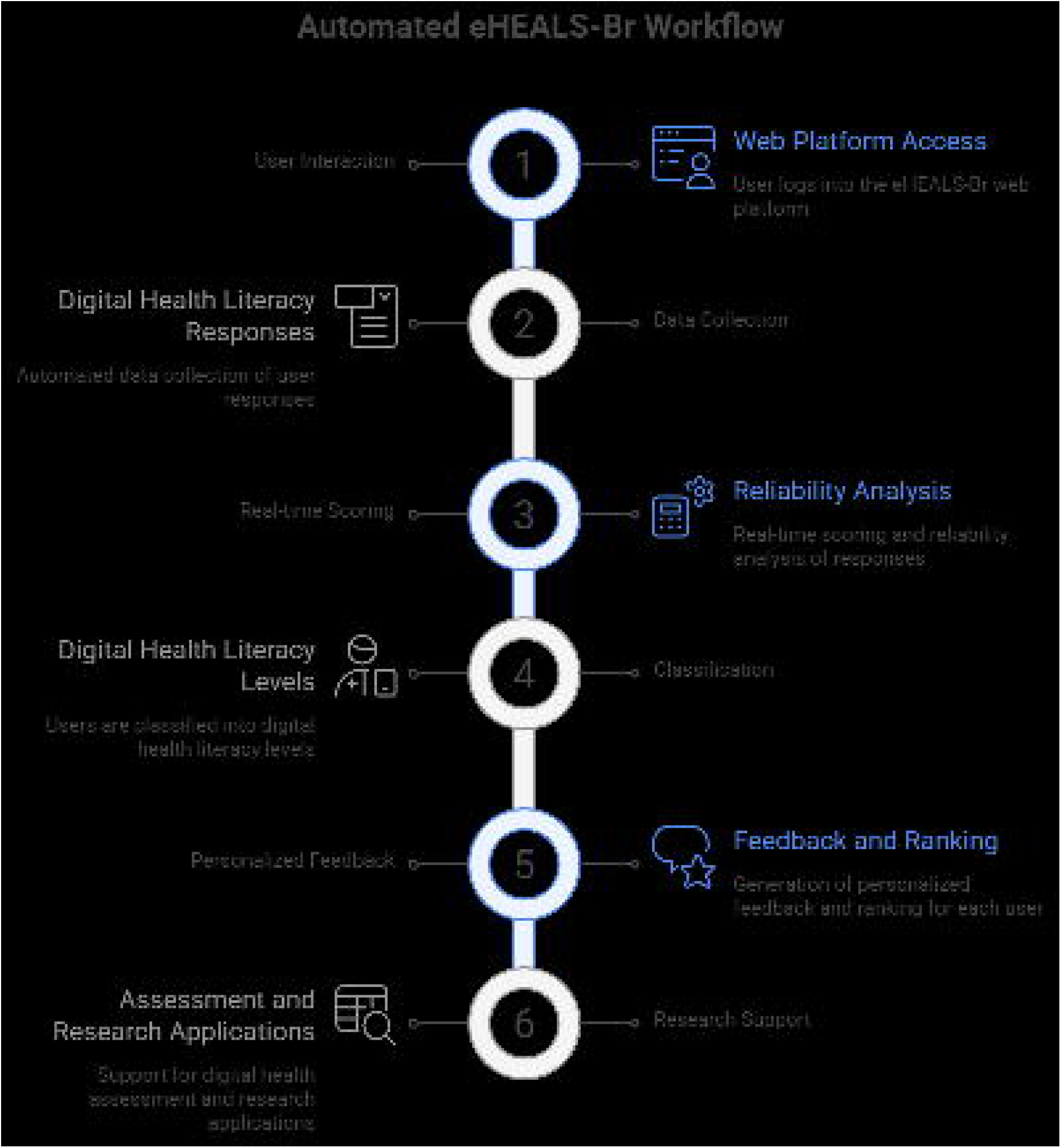

## Footnotes

1 Source: Haddaway et al. (2022) [16]. Used with the online tool available at https://estech.shinyapps.io/prisma_flowdiagram/, with data from the present study.

2 This software version was officially registered today with the Brazilian National Institute of Industrial Property (INPI) under **Process No. BR 51 2025 001179-2**, published in the *“RPI - Revista da Propriedade Industrial”*, Section VII - Computer Program. The record can be accessed at https://revistas.inpi.gov.br/rpi/. For direct consultation, the PDF of the issue is available at https://revistas.inpi.gov.br/pdf/Programa_de_computador2830.pdf; search for the authors’ names to locate the registration.

3 https://eheals.com.br

4 https://github.com/thallescotta/eheals.com.br

5 http://dgp.cnpq.br/dgp/espelhogrupo/9741699715904814; https://dippg.cefet-rj.br/ppgio/index.php/en/about-ppgio

